# RAPID POINT-OF-CARE DETECTION OF SARS-COV-2 BY ALVEO BE.WELL PLATFORM: ANALYTICAL AND CLINICAL EVALUATIONS

**DOI:** 10.1101/2024.03.07.24303877

**Authors:** Rixun Fang, Sha Coleman, Claudina Kwok, Helena Heisser, Teresa Wang, Lynn J. Kim, Ali Coopersmith, Darren Crawford, Kimberly Ha, Joseph C. Gaiteri, Yuh-Min (Johnson) Chiang

**Author notes:** **Corresponding author:**Rixun Fang Ph.D^1^.

## Abstract

**BACKGROUND:** The pandemic of coronavirus disease 2019 (COVID-19), caused by the severe acute respiratory syndrome coronavirus-2 (SARS-CoV-2), has had a profound global impact on human health and the economy. Early diagnosis and prompt effective treatment can reduce disease spread. At present, the majority of high-accuracy diagnostic tests for COVID-19 are conducted in specialized laboratories and rely on conventional quantitative real-time polymerase chain reaction (PCR) techniques. This diagnostic approach presents several challenges, including prolonged turnaround times and high costs due to the time and technical skills required to administer test protocols. Our primary aim is to develop a diagnostic platform suitable for use entirely within the point-of-care setting, avoiding the time and resource requirements of standard PCR test processing conducted away from the point-of-care.

**METHODS AND FINDINGS:** We developed the Alveo be.well COVID-19 Test as a diagnostic tool for qualitative detection of SARS-CoV-2 RNA in upper respiratory specimens. This innovative test detects viral RNA from SARS-CoV-2 within an unprocessed nasal specimen through the application of reverse transcription-loop mediated isothermal amplification (RT-LAMP) and electrical impedance measurement. The LAMP test primers are specifically designed to amplify a conserved region in the nucleocapsid gene of SARS-CoV-2 RNA, ensuring detection accuracy. To enhance usability and shelf life at ambient temperature, the primers are embedded within a microfluidic cartridge, along with other required reagents for viral target amplification. This test is designed for point-of-care settings, offering a straightforward and user-friendly process. The test includes: nasal swab sample collection; elution of the sample in a buffer; transfer of the eluted sample into the cartridge; insertion of the cartridge into an analyzer for amplification, and real-time result interpretation. All processing steps, including heating, mixing, amplification, and detection, occur within the cartridge during the test run, making it particularly user-friendly and obviating the need for significant user training. Results are displayed on a mobile smart device within approximately 50 minutes, via the be.well app, facilitating real-time decision-making in the point-of-care environment. Test and result data is also transferred to and stored in the cloud.

To evaluate the analytical performance of this platform, we initially assessed the analytical sensitivity and specificity of the Alveo be.well COVID-19 test. We also evaluated the diagnostic performance of the Alveo be.well COVID-19 assay using contrived reference samples. Furthermore, we validated the clinical performance of the Alveo be.well COVID-19 test in different clinical settings in both USA and United Kingdom during the pandemic seasons in 2020 through 2022. Test results from 253 nasal swab samples were compared to a real-time reverse transcription polymerase chain reaction (RT-PCR) reference standard.

**CONCLUSIONS:** The development of the Alveo be.well platform for infectious disease testing, including SARS-CoV-2 diagnosis, represents a significant advancement in portable diagnostic technology, encompassing both nucleic acid amplification and detection technologies. In testing SARS-CoV-2, the system amplifies the nucleic acid target within unprocessed nasal specimens using RT-LAMP chemistry, while concurrently measuring the electrical impedance signal produced during target amplification process, delivering results in under an hour. The system exhibits high analytical specificity and sensitivity, with a limit of detection of as low as 4000 viral genomic copies per milliliter, making it highly effective for detecting the SARS-CoV-2 target. Results derived from clinical validation studies showed 93% sensitivity and 95% specificity when compared to RT-qPCR. This system was demonstrated to be a user-friendly and rapid diagnostic platform with high analytical and clinical specificity and sensitivity for SARS-CoV-2 detection with the potential to facilitate early, fast diagnosis of SARS-CoV-2 to help contain the spread of the disease, particularly in point-of-care settings.

## INTRODUCTION

At the end of 2019 a new virus, SARS-CoV-2, emerged in Wuhan, China, causing a disease called COVID-19 [1]. The resulting COVID-19 pandemic has affected millions of people, resulting in significant loss of life and a global economic downturn. By September 2023, over 770 million individuals worldwide had tested positive for the virus, with more than 6.9 million lives tragically lost (2). The rapid identification and isolation of infected individuals has played a pivotal role in slowing the spread of the disease, particularly in point-of-care settings [1]. In response to the pandemic and rapid spread of SARS-CoV-2, a multitude of diagnostic tests have been developed. These tests encompass viral nucleic acid detection by PCR, antigen-based immunological tests for the virus, and serological tests for patients. It is important to note that SARS-CoV-2 patients are often infectious during the pre-symptomatic phase and may even never develop symptoms [2,3], so early detection is imperative in controlling the disease’s spread. Unfortunately, currently available tests have significant drawbacks. Antigen-based tests, while faster, often exhibit limitations in sensitivity, especially in detecting early-stage infections. This limitation can potentially lead to false negatives, particularly in cases where viral loads are low or during the initial phases of infection. Antibody tests are also flawed for early detection because IgG and IgM antibodies are generally undetectable until 7-10 days after exposure [4]. Viral loads reach their peak within the first days of the symptomatic period [2,4,5], making nucleic acid amplification tests the preferred method for detection during and prior to the symptomatic period. RT-PCR is the most widely used method for detecting RNA of SARS-CoV-2 [6]. Despite its high sensitivity RT-PCR does have its limitations for point-of-care diagnostics of COVID-19, which include its lack of portability and the need for specialized equipment and skilled personnel. PCR instruments are typically large and require precise temperature cycling and use complex optical detection methods [7,8]. Additionally, RT-PCR is poorly compatible with crude respiratory samples due to the presence of inhibitors [9] that necessitate extracting and concentrating the virus’s RNA, a process that either requires skilled technicians in a laboratory or a complex sample preparation apparatus. This makes RT-PCR less suitable for rapid testing at the point of care, such as in remote or resource-limited settings. Additionally, PCR may not be well-suited for situations where immediate results are needed, as the process can be time-consuming. To address these limitations, we have developed a portable and user-friendly device designed for point-of care COVID-19 testing. Our aim is to uphold the high sensitivity and specificity of RT-PCR while providing faster and accessible testing options, all without the need for sample preparation. In this paper, we present our developed portable diagnostic device, known as Alveo be.well™ platform (Figure 1), which has been analytically and clinically evaluated for the detection of SARS-CoV-2. As shown in Figure 1, the Alveo be.well system is composed of a nasal swab, be.well™ Assay Buffer, the be.well™ COVID-19 Cartridge, the be.well™ Analyzer and the be.well™ Mobile App. The be.well cartridge is an injection-molded plastic cassette with pathogen-specific amplification reagents dried within 8 reaction wells, each of which has electrodes to measure electrical impedance of the reaction solution.

**Figure 1.**
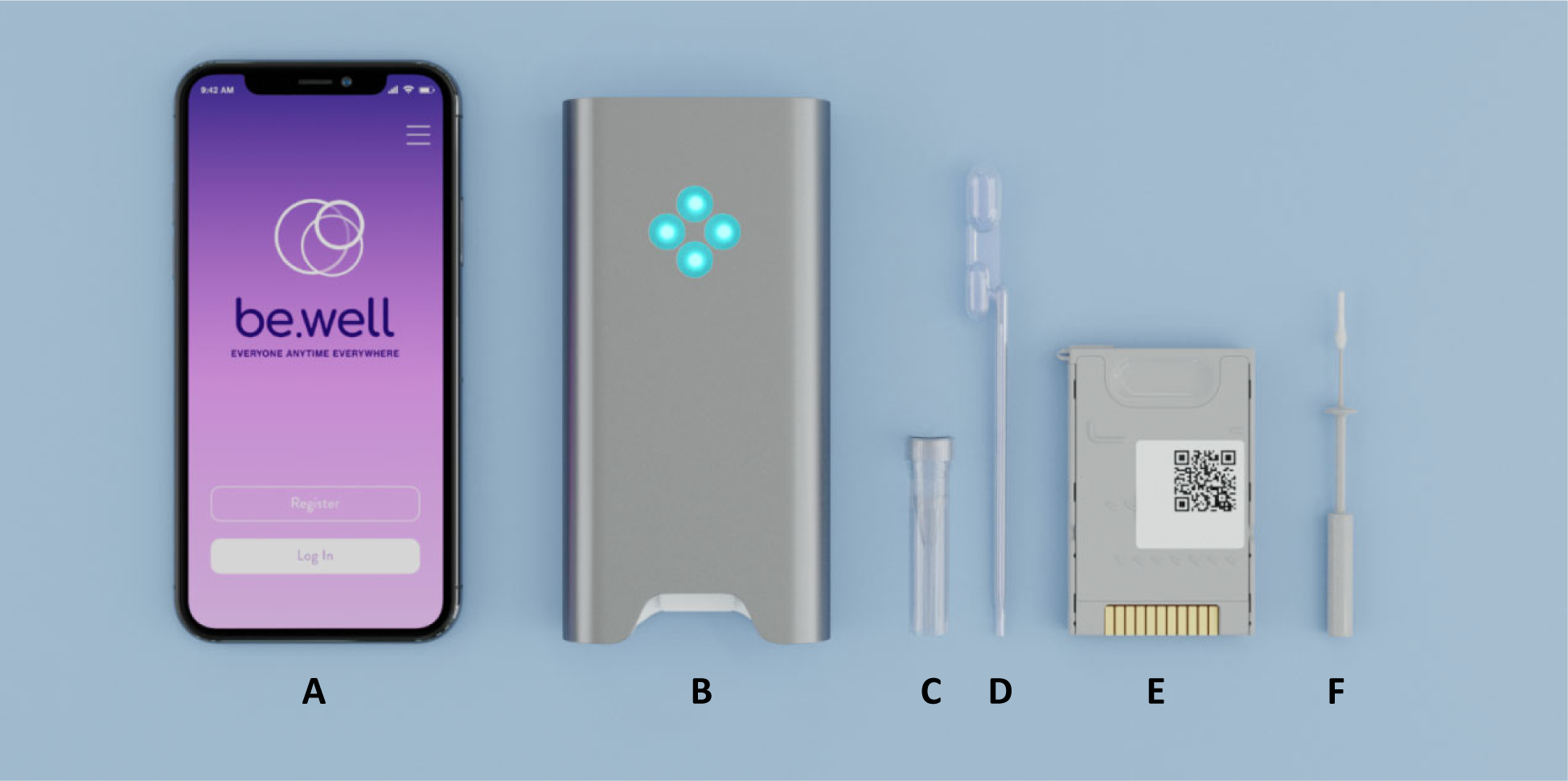
Alveo be.well^TM^ platform: (A) iPhone running be.well^TM^ software application (B) Analyzer (C) Assay buffer vial (D) Nasal swab (E) Disposable cartridge (F) Nasal swab

The platform is designed to use nasal swab samples (Fig. 2), commonly collected by a healthcare provider or patient using standard nasopharyngeal sampling techniques [10]. Swab samples from patients are eluted into an assay buffer vial and then loaded into the cartridge in a single pipetting step. Within the cartridge, the sample is simultaneously loaded into all wells via a microfluidic process. Each well can be configured to test distinct targets or controls, so each cartridge is capable of multiplexing up to eight reactions. Within the SARS-CoV-2 cartridge, two wells are dedicated to assay controls: one is a process control based on bacteriophage MS2, the other serine dehydratase, a housekeeping gene sample control. Four cartridge wells are dedicated to the SARS-CoV-2 assay for redundancy purposes. Once loaded, the cartridge is then inserted into the be.well Analyzer where the isothermal process is applied to induce amplification of the target.

**Figure 2.**
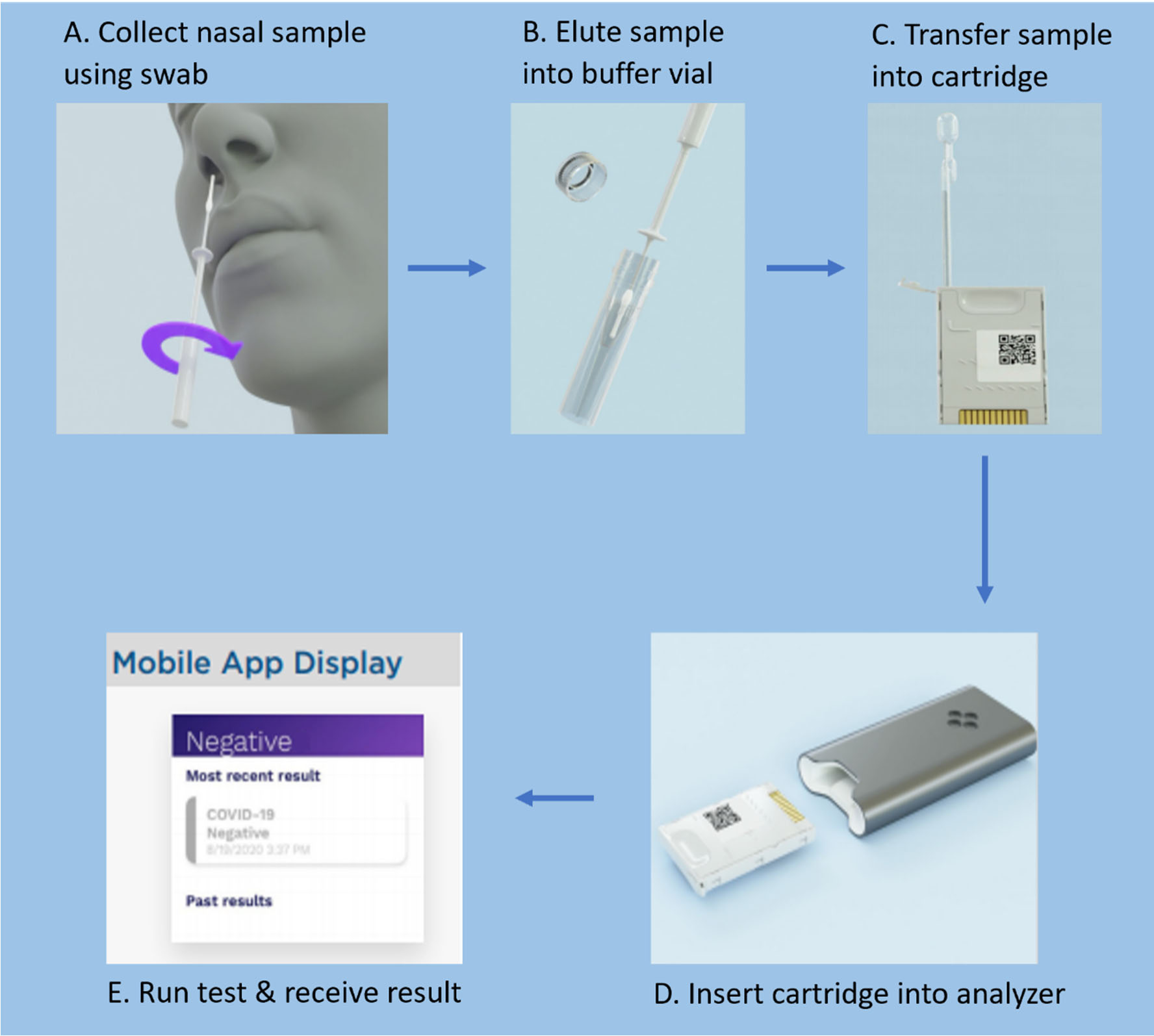
Assay workflow. (A) The sample is collected using a mid-turbinate nasal swab. (B) The sample is eluted into a vial containing assay buffer and mixed thoroughly. (C) The sample is transferred from the vial into the cartridge using either a transfer or precision pipette. This loads all eight wells within the cartridge. The cartridge cap is then closed to seal the contents. (D) The loaded cartridge is inserted into the be.well Analyzer, where the assay’s heating and detection are executed. (E) The be.well Mobile App is used to initiate the assay on the Analyzer. After the assay is completed, the App displays the result.

Our amplification chemistry is based on reverse-transcriptase loop-mediated isothermal amplification (RT-LAMP) [11,12], a technique with multiple characteristics that make it suitable for use in a compact, portable, self-contained device without requiring laboratory training or skills. RT-LAMP is isothermal, typically running at approximately 60-65 °C [12], removing requirements for temperature cycling. It is also compatible with crude matrices such as serum and nasal fluid, thus not requiring the typically time-consuming/labor-intensive isolation of viral RNA from a sample. In contrast to traditional real-time PCR fluorescence or turbidity detection, the Alveo be.well platform relies on the electrical properties of magnesium pyrophosphate produced by the amplification reaction. The be.well Analyzer monitors the reaction in real time by measuring the electrical impedance of the reaction solution within each well, which changes in response to the concentration of magnesium pyrophosphate (Figure 3). The result (COVID-19 Positive, COVID-19 Negative, or Invalid) is computed on a microcontroller within the analyzer and communicated to an iOS/Android app using a HIPAA-compliant Bluetooth connection. The electrical impedance sensing process is free from nucleotide fluorescence labeling and non-optical, minimizing instrumentation cost and complexity.

**Figure 3.**
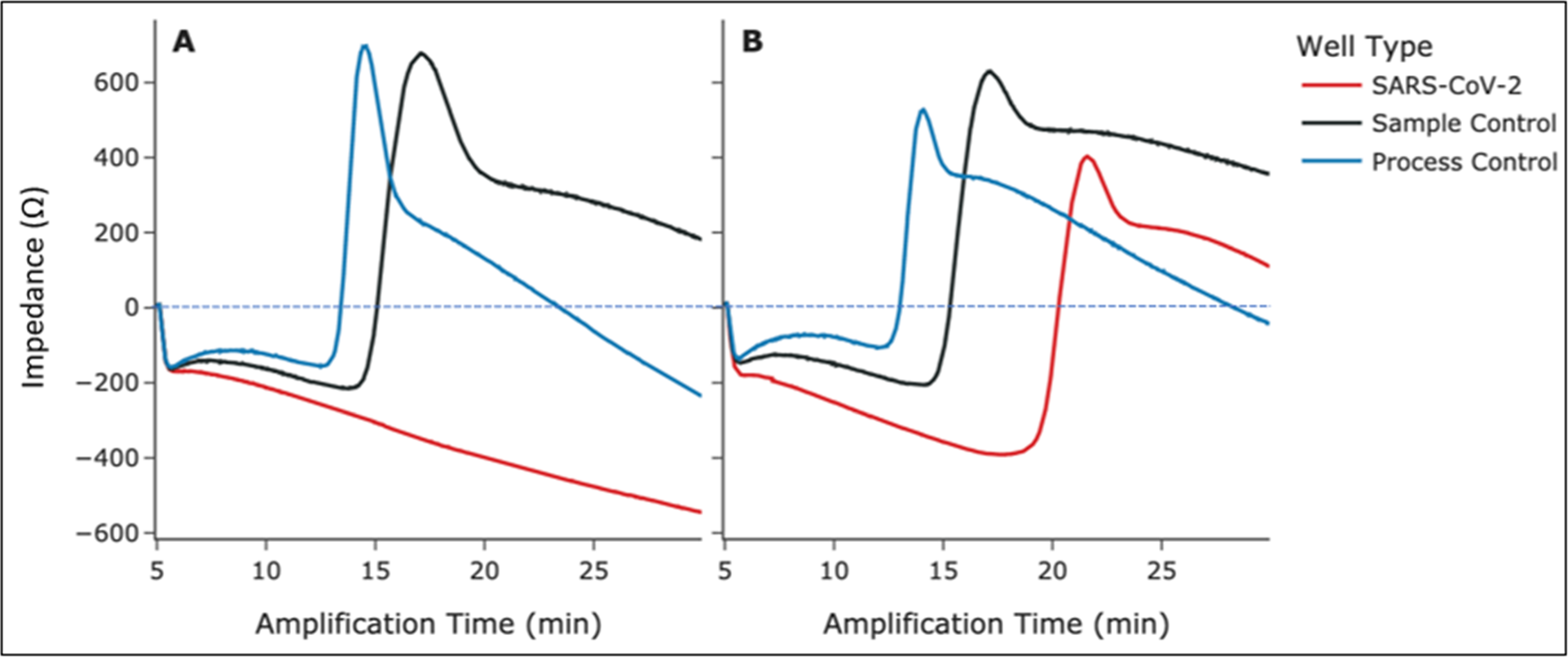
Amplification curves for (A) SARS-CoV-2 negative and (B) SARS-CoV-2 positive cartridges. Blue curves signify the process control, black curves signify the sample control, and red curves signify a SARS-CoV-2 well. All control wells amplified in both cartridges, while the SARS-CoV-2 well amplified in only the positive cartridge. The X axis represents the reaction run-time in minutes and the Y axis displays the detection signal, measured in Ohms as electrical impedance signal.

## METHODS AND MATERIALS

### ETHICAL STATEMENT

A total of 253 clinical samples (mid-turbinate swabs) were collected in USA and United Kingdom during the pandemic seasons in 2020, two clinical sites in the US (Florida and Texas) and six sites in the United Kingdom in both Hampshire, including Basingstoke and North Hampshire Hospital, Microbiology Laboratory, Royal Hampshire County Hospital, and Basingstoke Covid-19 Community Hub, as well as in London, which includes King’s College Hospital in Denmark Hill and the South London Specialist Virology Centre of KCH NHS Foundation Trust. The protocol used for collecting and detecting the patient samples was approved by relevant local Institutional Review Boards (IRBs).

### CHOICE OF SAMPLE

We designed our system for use with mid-turbinate nasal swabs (COPAN Diagnostics Inc.), as these can be collected by a healthcare provider or self-collected by patients. In analytical studies, all reactions contain pooled human nasal fluid (Lee Biosolutions) at 1% (v/v).

### PRIMER DESIGN

We downloaded a reference sequence for SARS-CoV-2’s nucleocapsid (N) gene from GenBank (accession number NC_045512.2, bp 28274-29533). Using MAFFT 7 [13] and Jalview v2.11 [14], we aligned the reference N gene to full genomic sequences downloaded from the GISAID Epi-CoV database [15] and parsed all unique N gene sequences from the alignment.

Based on the N gene alignment, we used proprietary in-house software written in Python 3 (Python Software Foundation) to generate primer sets for RT-LAMP. Sequences for all LAMP primer domains (F3/B3, F2/B2, LF/LB, F1/B1) were chosen based on their ability to maintain an optimal average melting temperature (Tm) against every SARS-CoV-2 strain while never falling below a minimum effective Tm for any given strain. Our software also screened each primer set *in silico* to minimize unwanted secondary structures such as primer dimers (strongest allowable dimer = −9.5 kcal/mol) and hairpins (maximum allowable hairpin Tm = 50 °C).

### OLIGONUCLEOTIDE AND TEMPLATE SEQUENCES

All primers were purchased from Integrated DNA Systems (IDT), as was a 700-bp double-stranded DNA target containing a fragment of the N gene sequence from GenBank Accession No. NC_045512.2. We used a MEGAscript T7 Kit (Thermo Fisher Scientific) to generate a 660-bp RNA transcript from the synthetic target. Heat-inactivated virus and gamma-inactivated virus (both SARS-CoV-2 isolate USA-WA1/2020) were obtained from BEI Resources. Genomic RNA (Hong Kong/VM20001061/2020) was obtained from American Type Culture Collection (ATCC).

### AMPLIFICATION PARAMETERS FOR PROTYPE ASSAY DEVELOPMENT

In the development of the protype SARS-CoV-2 RT-LAMP assay, liquid reactions were utilized and performed on a real-time PCR instrument. We used WarmStart LAMP Master Mix from New England Biolabs (NEB). Reactions contained 50 mM Na+, 8 mM Mg++, 5.6 mM total dNTPs, F3 and B3 primers at 0.2 µM each, LF and LB primers at 1 µM each, FIP and BIP primers at 1.6 µM each, 50 U murine RNase inhibitor (New England Biolabs), 4% Ficoll-400k (Sigma-Aldrich), and 1% human nasal fluid (Lee Biosystems). Reactions were run isothermally at 65 °C for up to 60 minutes. Initial primer screening reactions and cross-reactivity studies were performed on a QuantStudio 5 thermocycler (Thermo Fisher Scientific) using EvaGreen intercalating dye (Biotium) as a reporter. This enabled rapid throughput and, based on bridging studies performed in-house, is predictive of sample reactivity on our cartridge.

### CARTRIDGE STUDIES

A mixture of the reaction components, such as enzymes, dNTPs, and target-specific primers were dried directly within reaction wells on the cartridge. In the analytical limit of detection (LoD) studies, the equivalent of one swab (10 µL) of heat-inactivated SARS-CoV-2 (BEI) + nasal fluid (crude sample study) or purified genomic RNA (ATCC) + purified human DNA (Promega) were spiked into 500 µL Assay Buffer in 0.5-mL microcentrifuge vials. The mixed preparations were then pipetted into our cartridges and run in our analyzers using either 40 min at 65 °C (preliminary LoD) or 10 min at 50 °C followed by 40 min @ 65 °C (enhanced LoD study). Results were determined using either Internal Research and Investigative Software (in-house) or our be.well iOS app.

### CROSS-REACTIVITY STUDIES

In accordance with FDA recommendations, viral samples were tested at reaction concentrations of ≥100k PFU/mL, while bacterial samples were tested at ≥1 million CFU/mL. If viral samples were insufficiently concentrated, extraction + concentration procedures were performed with a PureLink Total RNA/DNA kit (Thermo Fisher Scientific) to ensure that sufficient quantities of the pathogen could be added to the reaction.

### IN SILICO ANALYSIS

We used in-house Python 3 scripts for all *in silico* analyses. For exclusivity studies, tables of k-mers for each primer length were generated from all target sequences, and the percentage of matching nucleotides (% homology) between each primer and equal-length k-mer were calculated. Reverse complements of each primer were also analyzed to account for homology in double-stranded templates. Any primer/k-mer combination exceeding 80% homology was flagged. The locations and sequences of high-homology k-mers were tracked to inspect potential amplicons.

### SOFTWARE AND DATABASES

Thermocycler data were analyzed using QuantStudio Design & Analysis Software v1.5.1 (Thermo Fisher), and cartridge run results were analyzed using in-house software written in C#. Data plots and tables were produced with JMP 15 (SAS Software, Inc.), Python 3, and Microsoft Excel.

## RESULTS

### PRIMER DESIGN

The SARS-CoV-2 assay targets a segment of the nucleocapsid (N) gene in accordance with WHO detection protocols. Primers were designed with a proprietary, internally developed RT-LAMP primer design software and an alignment of all available SARS-CoV-2 sequences from the GISAID EpiCoV database [15] as of March 17, 2020. Over 100,000 candidate primer sets were initially generated, from which eight were chosen for empirical testing in a thermal cycler. Purified genomic RNA of SARS-CoV-2 was used to determine an approximate limit of detection for the primer set in thermal cycler reactions. The best primers were chosen based on amplification speed (<20 minutes for most replicates), sensitivity, and lack of false-positive amplifications.

### PRIMER SPECIFICITY

To evaluate specificity, we screened our primer set *in silico* against a known panel of respiratory pathogens (Table 1), including common coronavirus strains (HKU1, NL63, OC43, 229E) and those most associated with severe outcomes (MERS and SARS). All instances of a primer showing ≥80% homology against any subsequence within a pathogen were flagged in accordance with FDA guidelines for *in silico* reactivity. While six bacterial species showed reactive homology against a single primer, this indicates a low risk for false-positive amplification due to the inability of a lone primer to amplify a target exponentially. Only SARS-CoV-1, a close genetic relative of SARS-CoV-2 (∼88% homology in the N gene), showed reactive homology against more than 1 primer.

**Table 1.** Pathogen genomes tested *in silico* against our primer set.

| **Pathogen Genomes Tested *In Silico* Against SARS-CoV-2 Primers** |  |
| --- | --- |
| **Viral Pathogens** | **Bacterial Pathogens** |
| Human coronavirus (229E, OC43, HKU1, NL63, MERS-CoV, SARS-CoV-1, SARS-CoV-2) | *Chlamydia pneumoniae* |
| Human adenovirus (1, 2, 3, 4, 5, 7, 14, 21) | *Haemophilus influenzae* |
| Influenza (A/H1N1, A/H1N1pdm09, A/H3N2, B) | *Legionella pneumophila* |
| Human metapneumovirus (A1, A2, B1, B2) | *Mycobacterium tuberculosis* |
| Human parainfluenza virus (1, 2, 3, 4a, 4b) | *Streptococcus pneumoniae* |
| Human respiratory syncytial virus (A, B) | *Streptococcus pyogenes* |
| Human rhinovirus (A, B, C) | *Bordetella pertussis* |
|  | *Mycoplasma pneumoniae* |
|  | *Pneumocystis jirovecii* |

Next, we tested a panel of these pathogens in wet reactions against our primer sets. In accordance with FDA guidelines, viral and bacterial samples were tested in the reactions at ≥100k PFU/mL and ≥1 million CFU/mL, respectively. No amplifications were seen within the 40-minute cutoff window for any of the pathogens, except for one *S. salivarius* replicate (∼35 min time to result, TTR) and the SARS-CoV-1 N gene plasmid. Due to the lack of homology between *S. salivarius* and our primers, we expect the lone amplification is the result of components in the bacterial broth. Mild cross-reactivity with the high-concentration (40 million C/mL) SARS-CoV-1 plasmid is likely the result of homology between SARS-CoV-1 and SARS-CoV-2 but we regard this as presenting no risk, as SARS-CoV-1 has not been identified in humans since 2004 [16]. All samples included 1% (v/v) human nasal fluid, which contains human DNA as well as genetic material from a diverse panel of respiratory microbes. The lack of amplification suggests that our assay will not cross-react with genetic material normally present in respiratory samples.

### INCLUSIVITY OF PRIMER SETS

To predict inclusivity, we used computer modeling of our primer set against a multiple sequence alignment (MSA) of SARS-CoV-2 genomes from the GISAID database [15] on December 30, 2023. We aligned each primer to each genome and calculated the respective Tm (melting temperature) using nearest-neighbor thermodynamic data from the SantaLucia lab [17]. A genome was considered detectable so long as all primers met predefined Tm thresholds. Of the >1.8 million sequences in the GISAID MSA, all met or exceeded the thresholds, indicating that our assay can detect all strains of SARS-CoV-2 currently in circulation. This includes highly infectious variants of concern such as Alpha, Delta, and Omicron, as their hallmark mutations are primarily in a region (the spike protein) that our assay does not target [18,19].

### LIMIT OF DETECTION STUDIES

#### Single-Step Heating LoD

We used simulated crude samples to conduct two limit of detection (LoD) studies with the Alveo be.well COVID-19 test. LoD is defined here as the lowest sample viral load that will be detected 95% of the time. In the first study, cartridge runs were performed for 40 min at 65°C, though positive results were determined in <25 min in most cases. To simulate unpurified samples assayed directly from patient swabs, varying concentrations of heat-inactivated SARS-CoV-2 virions were added to assay buffer containing 1% (v/v) human nasal fluid. These contrived samples were then loaded into cartridges and assayed. An initial 3-fold dilution series was performed with 3 replicates at each virus concentration. The highest concentration in the series without a 100% detection rate was 1,300 copies/sample (C/sample). We used the next highest concentration in the series (4,000 C/sample) as a starting point for a series of increasingly concentrated dilutions at up to 20 replicates each. Beginning our increasing series at 4,000 C/sample, detection rates at each concentration were found to be between 70% and 86% until we reached a 100% detection rate at 17,000 C/sample (Figure 4).

**Figure 4.**
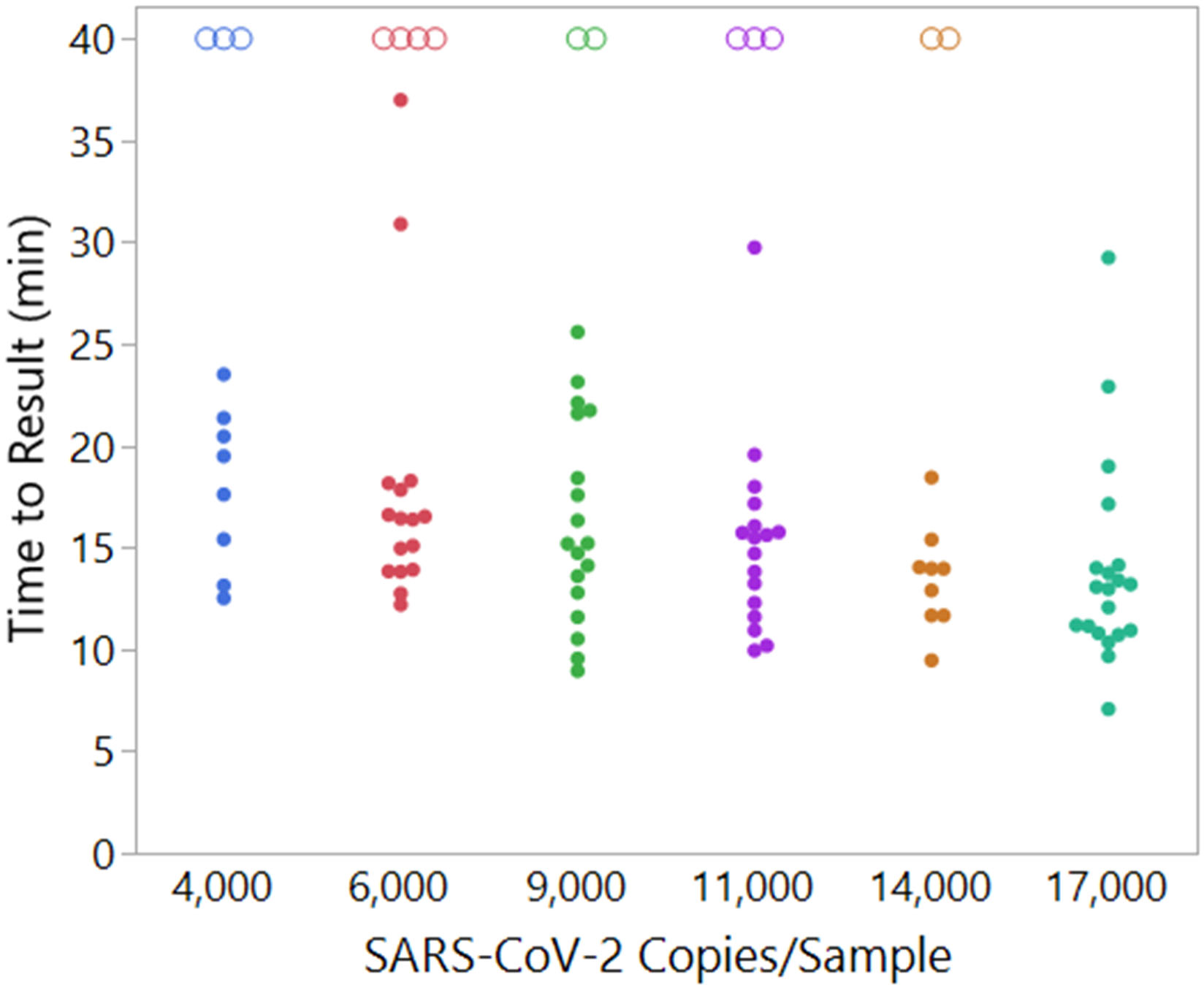
Result times for each cartridge in the single-step heating LoD study. Each point represents the time to result from one cartridge. Empty circles at 40 min are placeholders for cartridges that failed to amplify. 17,000 copies/sample is the lowest concentration at which ≥95% of cartridges amplified. Average time to result was 14.1 min.

#### Multi-Step Heating LoD

The second LoD study was designed to improve rehydration of dried reagents and increase the efficiency of the reverse transcription step before proceeding to exponential DNA amplification. In this study, cartridges were held for 10 min at ambient conditions (approx. 21 °C) and then for 10 minutes at 50 °C before increasing the temperature to 65 °C for LAMP. Prior to testing positive samples, 20 blank cartridges were run to determine the appropriate cutoff time for the assay. Using 5 cartridges per concentration, we ran a twofold dilution series beginning with 8,000 C/sample down to 250 C/sample. We achieved 100% detection (5 out of 5 cartridges) at all concentrations except for 250 C/sample, where 3 out of 5 cartridges detected SARS-CoV-2 virus. To determine the actual LoD, we then ran 500 C/sample and 2,000 C/sample at 20+ cartridges per concentration. Using an assay cutoff time of 30 min at 65 °C, we achieved a hit rate of 85% at 500 copies/sample (equivalent to 1,000 C/mL) and 95% at 2,000 copies/sample (equivalent to 4,000 C/mL) (Figure 5). If our 65 °C cutoff time were 20 min instead of 30 min, we would still detect 95% of cartridges at 2,000 C/sample using the same assay run time as in the initial, one-step LoD study. This value corresponds to 4,000 C/mL in the buffer tube and implies that our system can detect approximately 100 copies in each 25 µL reaction well (see Figure 5) 95% of the time. This LoD is on the same order as that of the CDC’s EUA-authorized RT-PCR kit (LoD = 1,000 C/mL) [20] and without requiring sample extraction or a thermocycler.

**Figure 5.**
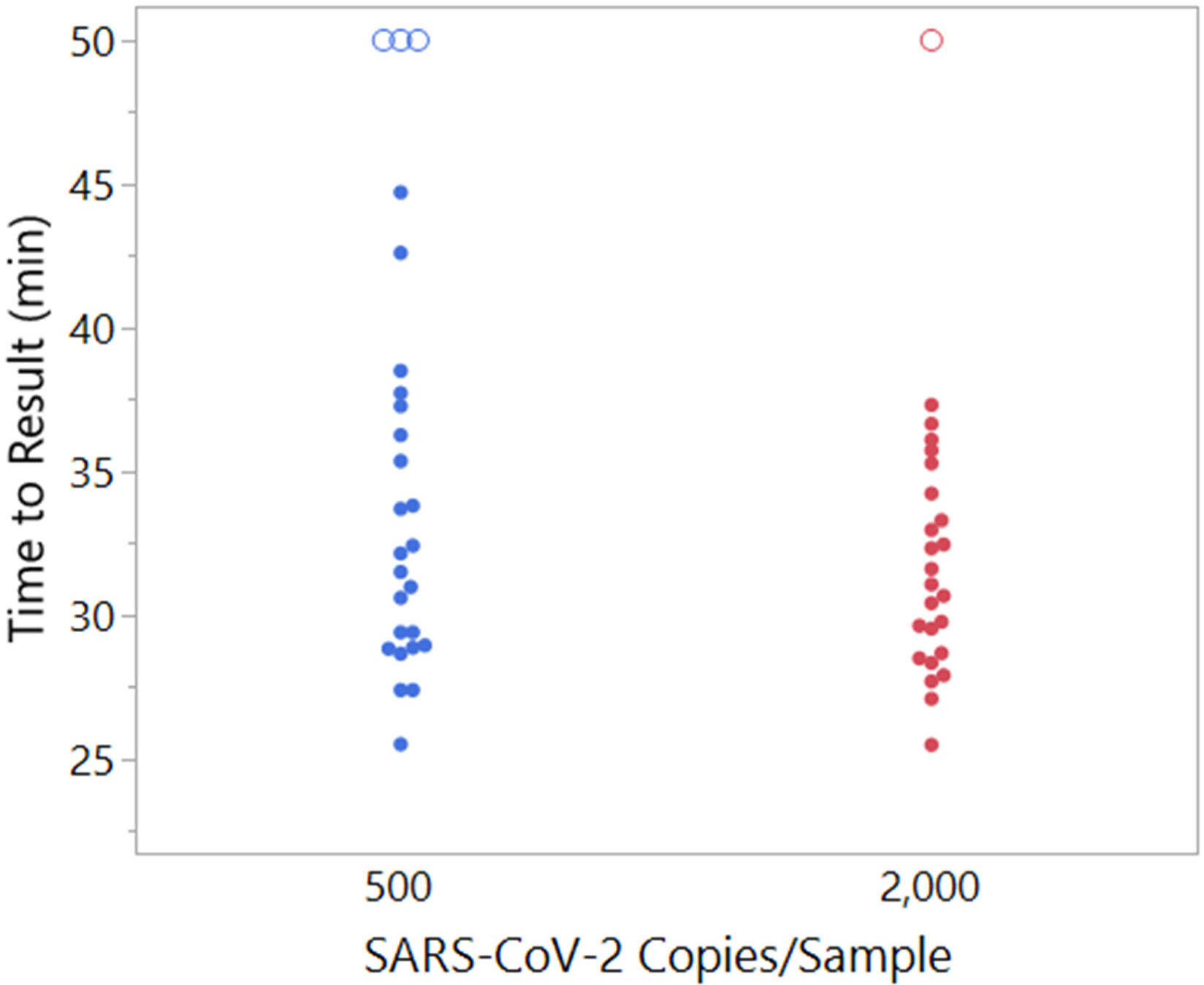
Result times for each cartridge in the multi-step heating LoD study. Each point represents the time to result from one cartridge. Empty circles at 50 min are placeholders for cartridges that failed to amplify. 2,000 copies/sample is the lowest concentration at which ≥95% of cartridges amplified. Average time to result was 31.4 min. These are clinically significant concentrations owing to patients shedding below approximately 100,000 copies/swab generally being noninfectious [2,21].

### CLINICAL EVALUATION

To evaluate the clinical performance of the be.well COVID-19 assay, two IRB-approved prospective clinical trials were conducted during September 2020 through December 2020, with one trial taking place in the United States and the other in the United Kingdom. For the US clinical evaluation study, patient samples were collected from two clinical sites (Florida and Texas) using the Copan mid-turbinate swabs for both Alveo be.well COVID-19 test and comparator PCR test authorized by FDA. Both samples were shipped to a testing site in California after collection and tested with the be.well COVID-19 test and the EUA authorized Abbott RealTime SARS-CoV-2 Assay performed on the Abbott m2000 System. The be.well COVID-19 test results were compared to results from the Abbott RealTime SARS-CoV-2 Assay to determine the clinical sensitivity and specificity and the 95% confidence interval (CI). The positive percent agreement (sensitivity) was calculated to be 94% (95% CI, 79.2% to 99.2%) with a negative percent agreement (specificity) of 97% (95% CI, 92.7% to 99.2%) as shown in Table 2.

**Table 2.** Performance of the be.well COVID-19 test for detection of SARS-CoV-2 virus in comparison with Real Time qPCR based SARS-CoV-2 Assays.

|  | TP | FP | TN | FN | Total | % Sensitivity<br>(95% CI) | % Specificity<br>(95% CI) |
| --- | --- | --- | --- | --- | --- | --- | --- |
| USA | 30 | 2 | 133 | 4 | 179 | 94%<br>(79.2% - 99.2%) | 97%<br>(92.7% - 99.2%) |
| UK | 28 | 2 | 42 | 2 | 74 | 93%<br>(77.9% - 99.2%) | 95%<br>(84.5% - 99.4%) |
| Combined | 58 | 4 | 175 | 6 | 253 | 94%<br>(84.6% - 97.5%) | 95%<br>(93.2% - 98.4%) |

FN: false-negative; FP: false-positive; TN: true-negative; TP: true-positive.

The prospective clinical performance evaluation was also carried out in multiple sites in Hampshire and London of the United Kingdom. Testing was spread across four sites in Hampshire (Basingstoke and North Hampshire Hospitals, Microbiology Laboratory, Basingstoke and North Hampshire Hospitals) and 2 sites in London (King’s College Hospital and Denmark Hill London). Two samples were collected from all the patients. The first sample was taken from the nostril of the patient using Copan pediatric flocked swabs for the Alveo be.well Test according to the instructions of the assay. The second sample was collected from the nose and throat using a nasopharyngeal adult swab and preserved in viral transport media. The first sample was then tested immediately in the be.well COVID-19 test on site, while the second sample was taken to the local laboratory for PCR testing. The Altona RealStar® SARS-CoV-2 RT-PCR Kit 1.0 with an LOD of 100 copies/mL was used as the comparator instruments in London, while the CERTEST VIASURE SARS-CoV-2 Real Time PCR Detection Kit with an LOD of 20 copies/reaction was used in Hampshire. After the study, the PCR results were combined from both London and Hampshire sites for comparison to the be.well COVID-19 results. Ct values for positive specimens in the Abbott assay ranged from 16-45. The combined clinical sensitivity (positive percent agreement) and specificity (a negative percent agreement) was demonstrated to be 94% (95% CI, 84.6% - 97.5%) and 95% (95% CI, 93.2%-98.4%), respectively (Table 2).

## DISCUSSION

Viral load estimates in the literature typically report median values on the order of 10,000 C/mL to 1 million C/mL or higher at time of diagnosis, depending on factors such as patient age, disease severity, and days since symptom presentation [2]. As such, we expect that an LoD of approximately 4,000 C/mL (∼2,000 C/sample in our system) will be sufficient to confirm either mild or severe illness. Furthermore, because infectivity (measured by successful viral culture from patient samples) appears highly unlikely below 10^5^–10^6^ C/sample [2,21], our system should identify the vast majority of infectious individuals.

We have demonstrated that the Alveo be.well platform can rapidly (under an hour time-to-result) and accurately detect SARS-CoV-2 using an innovative electrical sensing technology coupled with an isothermal amplification viral RNA detection chemistry. It differs from most current platforms in its ability to use raw patient samples from nasal swabs that can be collected and used for viral detection without utilizing complicated, sometimes scarce nucleic acid extraction kits. When these crude samples are used, our assay’s limit of detection is up to 2 orders of magnitude below the median viral load reported in many studies [2].

In conclusion, the be.well COVID-19 test is a new, simple, easy-to-use, and cost-effective RT-LAMP based nucleic acid detection platform for the fast and accurate detection of SARS-CoV-2. It is highly sensitive, with an LOD of 4,000 copies of SARS-CoV-2 viral RNA per mL, and it is specific regarding common respiratory pathogens. Clinical validation using over 200 patient samples in multiple clinical settings and sites demonstrates the robustness of the system and further confirmed that the assay’s performance was comparable to that of the RT-qPCR assay in the detection of SARS-CoV-2. This novel SARS-CoV-2 RT-LAMP detection platform overcomes the limitation of RT-qPCR assay and will become a powerful tool for SARS-CoV-2 detection and can be used to monitor the SARS CoV-2 virus in real time in all different types of clinical settings, which will be especially useful in COVID-19 surveillance and prevention of the spread of the disease.

## Data Availability

All data produced in the present work are contained in the manuscript.

## ACKNOWLEDGMENTS

UW, Monogram, UK, Don Green (wrote data analysis software), other people at Alveo for designing/manufacturing cartridges.

## REFERENCES

1. Larremore DB, Wilder B, Lester E, Shehata S, Burke JM, Hay JA, et al. Test sensitivity is secondary to frequency and turnaround time for COVID-19 surveillance. Infectious Diseases (except HIV/AIDS); 2020 Jun. doi:10.1101/2020.06.22.20136309

2. Jones TC, Biele G, Mühlemann B, Veith T, Schneider J, Beheim-Schwarzbach J, et al. Estimating infectiousness throughout SARS-CoV-2 infection course. Science. 2021;373: eabi5273. doi:10.1126/science.abi5273

3. Wang C, Horby PW, Hayden FG, Gao GF. A novel coronavirus outbreak of global health concern. The Lancet. 2020;395: 470–473. doi:10.1016/S0140-6736(20)30185-9

4. To KK-W, Tsang OT-Y, Leung W-S, Tam AR, Wu T-C, Lung DC, et al. Temporal profiles of viral load in posterior oropharyngeal saliva samples and serum antibody responses during infection by SARS-CoV-2: an observational cohort study. The Lancet Infectious Diseases. 2020;20: 565–574. doi:10.1016/S1473-3099(20)30196-1

5. Pan Y, Zhang D, Yang P, Poon LLM, Wang Q. Viral load of SARS-CoV-2 in clinical samples. The Lancet Infectious Diseases. 2020;20: 411–412. doi:10.1016/S1473-3099(20)30113-4

6. Service R. The standard coronavirus test, if available, works well—but can new diagnostics help in this pandemic? Science. 2020 [cited 11 Aug 2021]. doi:10.1126/science.abb8400

7. Ahrberg CD, Manz A, Chung BG. Polymerase chain reaction in microfluidic devices. Lab Chip. 2016;16: 3866–3884. doi:10.1039/C6LC00984K

8. Jauset-Rubio M, Svobodová M, Mairal T, McNeil C, Keegan N, Saeed A, et al. Ultrasensitive, rapid and inexpensive detection of DNA using paper based lateral flow assay. Sci Rep. 2016;6: 37732. doi:10.1038/srep37732

9. Schrader C, Schielke A, Ellerbroek L, Johne R. PCR inhibitors - occurrence, properties and removal. J Appl Microbiol. 2012;113: 1014–1026. doi:10.1111/j.1365-2672.2012.05384.x

10. Tu Y-P, Jennings R, Hart B, Cangelosi GA, Wood RC, Wehber K, et al. Swabs Collected by Patients or Health Care Workers for SARS-CoV-2 Testing. N Engl J Med. 2020;383: 494–496. doi:10.1056/NEJMc2016321

11. Nagamine K, Hase T, Notomi T. Accelerated reaction by loop-mediated isothermal amplification using loop primers. Molecular and Cellular Probes. 2002;16: 223–229. doi:10.1006/mcpr.2002.0415

12. Notomi T. Loop-mediated isothermal amplification of DNA. Nucleic Acids Research. 2000;28: 63e–663. doi:10.1093/nar/28.12.e63

13. Katoh K, Rozewicki J, Yamada KD. MAFFT online service: multiple sequence alignment, interactive sequence choice and visualization. Briefings in Bioinformatics. 2019;20: 1160–1166. doi:10.1093/bib/bbx108

14. Waterhouse AM, Procter JB, Martin DMA, Clamp M, Barton GJ. Jalview Version 2--a multiple sequence alignment editor and analysis workbench. Bioinformatics. 2009;25: 1189–1191. doi:10.1093/bioinformatics/btp033

15. Elbe S, Buckland-Merrett G. Data, disease and diplomacy: GISAID’s innovative contribution to global health: Data, Disease and Diplomacy. Global Challenges. 2017;1: 33–46. doi:10.1002/gch2.1018

16. About Severe Acute Respiratory Syndrome (SARS). In: Centers for Disease Control and Prevention [Internet]. [cited 11 Aug 2021]. Available: https://www.cdc.gov/sars/about/index.html

17. SantaLucia J, Hicks D. The Thermodynamics of DNA Structural Motifs. Annu Rev Biophys Biomol Struct. 2004;33: 415–440. doi:10.1146/annurev.biophys.32.110601.141800

18. Khateeb J, Li Y, Zhang H. Emerging SARS-CoV-2 variants of concern and potential intervention approaches. Crit Care. 2021;25: 244. doi:10.1186/s13054-021-03662-x

19. Gobeil SM-C, Janowska K, McDowell S, Mansouri K, Parks R, Stalls V, et al. Effect of natural mutations of SARS-CoV-2 on spike structure, conformation, and antigenicity. Science. 2021;373: eabi6226. doi:10.1126/science.abi6226

20. CDC 2019-Novel Coronavirus (2019-nCoV) Real-Time RT-PCR Diagnostic Panel. 2020. Available: https://www.fda.gov/media/134922/download

21. Wölfel R, Corman VM, Guggemos W, Seilmaier M, Zange S, Müller MA, et al. Virological assessment of hospitalized patients with COVID-2019. Nature. 2020;581: 465–469. doi:10.1038/s41586-020-2196-x

